# Kidney impairment is associated with in-hospital death of COVID-19 patients

**DOI:** 10.1101/2020.02.18.20023242

**Authors:** Yichun Cheng, Ran Luo, Kun Wang, Meng Zhang, Zhixiang Wang, Lei Dong, Junhua Li, Ying Yao, Shuwang Ge, Gang Xu

## Abstract

**Background:** Information on kidney impairment in patients with coronavirus disease 2019 (COVID-19) is limited. This study aims to assess the prevalence and impact of abnormal urine analysis and kidney dysfunction in hospitalized COVID-19 patients in Wuhan.

**Methods:** We conducted a consecutive cohort study of COVID-19 patients admitted in a tertiary teaching hospital with 3 branches following a major outbreak in Wuhan in 2020. Hematuria, proteinuria, serum creatinine concentration and other clinical parameters were extracted from the electronic hospitalization databases and laboratory databases. Incidence rate for acute kidney injury (AKI) was examined during the study period. Association between kidney impairment and in-hospital death was analyzed.

**Results:** We included 710 consecutive COVID-19 patients, 89 (12.3%) of whom died in hospital. The median age of the patients was 63 years (inter quartile range, 51-71), including 374 men and 336 women. On admission, 44% of patients have proteinuria hematuria and 26.9% have hematuria, and the prevalence of elevated serum creatinine and blood urea nitrogen were 15.5% and 14.1% respectively. During the study period, AKI occurred in 3.2% patients. Kaplan–Meier analysis demonstrated that patients with kidney impairment have higher risk for in-hospital death. Cox proportional hazard regression confirmed that elevated serum creatinine, elevated urea nitrogen, AKI, proteinuria and hematuria was an independent risk factor for in-hospital death after adjusting for age, sex, disease severity, leukocyte count and lymphocyte count.

**Conclusions:** The prevalence of kidney impairment (hematuria, proteinuria and kidney dysfunction) in hospitalized COVID-19 patients was high. After adjustment for confounders, kidney impairment indicators were associated with higher risk of in-hospital death. Clinicians should increase their awareness of kidney impairment in hospitalized COVID-19 patients.

## Introduction

In December 2019, a series of unknown origins cases of acute respiratory illness occurred in Wuhan, Hubei Province, China^1-2^. High-throughput sequencing indicated a novel betacoronavirus that is currently named “severe acute respiratory syndrome coronavirus 2” (SARS-CoV-2)^3^. On February 11^th^ 2020 the World Health Organization (WHO) officially named disease caused by SARS-CoV-2 as “Coronavirus Disease 2019” (COVID-19). The disease has rapidly spread from Wuhan to other areas globally. As of February, 17th, Chinese health authorities announced that 70641 confirmed cases of novel coronavirus infection and 1772 death cases had been reported in 31 provincial-level regions. Of note, in Wuhan, 41152 COVID-19 cases with 1309 deaths were confirmed at the same day, which indicated that the proportion of severe cases and the mortality rate in Wuhan were much higher than those in other provinces in China. However, the clinical characteristics of COVID-19 cases remain largely unclear. Identifying and eliminating factors that predict a negative outcome is the key to improve survival from COVID-19, especially in Wuhan.

Although diffuse alveolar damage and acute respiratory failure were the main features of COVID-19^4^, the involvement of other organs need to be considered. After lung infection, the infiltrated virus may enter the blood circulation, accumulate in kidney and cause damage to renal resident cells. Indeed, RNAaemia, defined as a positive result for real-time PCR in the plasma sample, was found in 15% COVID-19 patients^4^. It is reported that 6.7% patients with severe acute respiratory syndrome (SARS) in 2003 developed acute renal impairment and the mortality of SARS patients with acute kidney injury (AKI) was 91.7%^5^. Thus, the kidney impairment and outcome in patients infected by SARS-CoV-2, which resembles SARS in 2003, were urgently warranted.

In this large consecutive cohort study of COVID-19 adult patients in a tertiary teaching hospital with 3 branches and more than 4000 beds, which was designated for critical COVID-19 cases by local government, we aimed to demonstrate the prevalence and in-hospital outcome of kidney impairments in COVID-19 patients.

## Methods

### Participants

All consecutive COVID-19 patients admitted to Tongji hospital, Tongji medical college, Huazhong university of science and technology from January 28 to February 11, 2020 were enrolled. Tongji hospital, located in Wuhan, Hubei Province, the endemic areas of COVID-19, is one of the major tertiary teaching hospitals. Tongji hospital was assigned responsibility for the treatments of severe COVID-19 patients by Wuhan government on January 31^th^. All patients who were enrolled in this study were diagnosed as COVID-19 according to the guidance provided by the Chinese National Health Commission. Patients with a history of maintenance dialysis or renal transplantation were excluded. The clinical outcomes were monitored up to February 17, 2020, the final date of follow-up.

### Data Sources

The epidemiological characteristics, clinical symptoms and laboratory data were extracted from electronic medical records. Laboratory data consisted of complete blood count, liver and renal function, coagulation function, high-sensitivity C-reactive protein, procalcitonin, erythrocyte sedimentation rate, lactate dehydrogenase and creatine kinase. Estimated glomerular filtration rate (eGFR) was calculated with Chronic Kidney Disease Epidemiology Collaboration (CKD-EPI) equation^6^. The data were reviewed by a trained team of physicians. The date of disease onset was defined as the day when the symptom was noticed. The endpoint was the in-hospital death.

### Definition

Severity of the disease was staged according to the guidelines for diagnosis and treatment of COVID-19 (trial fifth edition) published by Chinese National Health Commission on February 4, 2020. Severe case was defined as either: (i) respiratory rate > 30/min, or (ii) oxygen saturation ≤ 93%, or (iii) PaO2/FiO2 ratio ≤ 300mmHg. Critical severe case was defined as including one criterion as follow: shock; respiratory failure requiring mechanical ventilation; combined with the other organ failure admission to intensive care unit (ICU).

AKI was defined as an increase in serum creatinine (Scr) by 0.3 mg/dL within 48 hours or a 50% increase in Scr from the baseline within 7 days according to the Kidney Disease: Improving Global Outcomes (KDIGO) criteria^7^. Baseline Scr was defined as the Scr value on admission. The date of AKI onset was defined as the earliest day that the Scr change met the KDIGO criteria. The stage of AKI was determined using the peak Scr level after AKI detection, with increase 1.5-1.9, 2.0-2.9 and ≥ 3 times baseline being defined as stage 1, 2 and 3, respectively.

### Statistical Analysis

Categorical variables were summarized as percentages, and continuous variables were expressed as the mean ± standard deviation or median with interquartile range. two-sample t tests or Wilcoxon rank-sum tests was used for continuous variables, and Chi-square tests or Fisher’s exact tests were used for categorical variables as appropriate. The candidate risk factors included age, sex, the severity of COVID-19 and laboratory data. Cumulative rates of in-hospital death were determined using the Kaplan–Meier method. The risk factors and corresponding HRs were calculated using the Cox proportional hazard model. Statistical analyses were performed using R software, version 3.6.1, with statistical significance set at 2-sided P<0.05.

## Results

### Baseline characteristics

A total of 710 patients were included in our study. The median age was 63 (51-71) years, and 52.7% were males. The median duration from illness onset to admission was 10 (7-13) days (Table 1). The mean level of lymphocyte count was 0.9 ± 0.5×10^9^/L, which were below the normal level. Most patients demonstrated elevated levels of high-sensitive C-reactive protein (83.1%) and erythrocyte sedimentation rate (81.8%), but elevated levels of procalcitonin were rare (10.7%). The coagulant function abnormality was common in COVID-19 patients. In addition, the mean level of lactose dehydrogenase (378 ± 195 U/L) was much higher than the normal level (Table 2).

**Table 1.** Baseline characteristics of patients with COVID-2019 Patients.

| Variables | All patients | Normal baseline serum creatinine | Elevated baseline serum creatinine | P-value |
| --- | --- | --- | --- | --- |
| Number | 710 | 600 | 110 |  |
| Age, years | 63 (51-71) | 61 (49-69) | 72 (62-79) | <0.001 |
| Male patients, % | 374/710 (52.7) | 294/600 (49.0) | 80/110 (72.7) | <0.001 |
| Days from illness onset to admission, days | 10 (7-13) | 10 (7-13) | 10 (7-12) | 0.511 |
| Fever on admission, % | 216/664 (32.5) | 187/560 (33.4) | 29/104 (27.9) | 0.324 |
| Respiratory rate > 30/min, % | 49/693 (7.1) | 39/585 (6.7) | 10/108 (9.3) | 0.446 |
| Systolic blood pressure, mmHg | 128 (117-143) | 128 (117-142) | 129 (116-145) | 0.454 |
| Diastolic blood pressure, mmHg | 79 (72-87) | 79 (72-87) | 77 (70-87) | 0.223 |
| Severe disease, % | 252/710 (35.5) | 198/600 (33.0) | 54/110 (49.0) | <0.001 |
| Acute kidney injury, % | 22/710 (3.2) | 12/600 (2.0) | 10/110 (9.1) | <0.001 |
| Stage 1 | 8/710 (1.3) | 7/600 (1.2) | 1/110 (0.9) | 0.036 |
| Stage 2 | 6/710 (0.8) | 1/600 (0.2) | 5/110 (4.5) |  |
| Stage 3 | 8/710 (1.1) | 4/600 (0.7) | 4/110 (3.6) |  |
| Days from admission to acute kidney injury, days | 4 (2-7.5) | 6 (4-8) | 2 (1.5-4.5) | 0.084 |
| In-hospital death, % | 89/710 (12.5) | 55 (9.2) | 34 (30.9) | <0.001 |

Data are presented as number/total (percentage) or median (interquartile range); COVID-2019, coronavirus disease 2019; The severity was staged based on the guidelines for diagnosis and treatment of COVID-19 (trial fifth edition) published by Chinese National Health Commission in February 4, 2020.

**Table 2.** Baseline laboratory data of patients with COVID-19 Patients.

| Variables | All patients | Normal baseline serum creatinine | Elevated baseline serum creatinine | P-value |
| --- | --- | --- | --- | --- |
| Leukocyte count, $\times 10^9/L$ | $7.5 \pm 7.5$ | $7.2 \pm 7.4$ | $9.5 \pm 7.8$ | 0.003 |
| Lymphocyte count, $\times 10^9/L$ | $0.9 \pm 0.5$ | $0.9 \pm 0.5$ | $0.8 \pm 0.5$ | 0.003 |
| Hemoglobin, g/L | $128 \pm 18$ | $127 \pm 17$ | $129 \pm 22$ | 0.498 |
| Platelet count, $\times 10^9/L$ | $212 \pm 94$ | $216 \pm 94$ | $190 \pm 93$ | 0.008 |
| Prothrombin time > 14.5s, % | 264/678 (38.9) | 215/573 (37.5) | 49/105 (46.7) | 0.097 |
| Activated partial thromboplastin time > 42s, % | 216/503 (42.9) | 171/423 (40.4) | 45/80(56.3) | 0.012 |
| D-dimer > 0.5 mg/L, % | 520/669 (77.7) | 139/424 75.3 | 96/106 (90.6) | 0.001 |
| Procalcitonin $\geq 0.5$ng/mL, % | 67/627 (10.7) | 37/538 (6.9) | 30/89 (33.7) | <0.001 |
| High-sensitivity C-reactive protein $\geq 10$mg/L, % | 567/682 (83.1) | 478/581 (88.1) | 89/101 (82.2) | 0.192 |
| Erythrocyte sedimentation rate > 15mm/h, % | 549/671 (81.8) | 463/568 (81.5) | 86/103 (83.5) | 0.733 |
| Alanine aminotransferase, U/L | $36 \pm 41$ | $36 \pm 40$ | $34 \pm 47$ | 0.021 |
| Aspartate aminotransferase, U/L | $43 \pm 50$ | $41 \pm 43$ | $53 \pm 77$ | <0.001 |
| Total bilirubin, mmol/L | $12 \pm 23$ | $11 \pm 7$ | $21 \pm 55$ | 0.060 |
| Lactose dehydrogenase, U/L | $378 \pm 195$ | $364 \pm 180$ | $458 \pm 248$ | <0.001 |
| Creatinine kinase, U/L | $165 \pm 232$ | $149 \pm 185$ | $252 \pm 388$ | 0.083 |
| Sodium, mmol/L | $139 \pm 5$ | $138 \pm 5$ | $139 \pm 7$ | 0.258 |
| Potassium, mmol/L | $4.2 \pm 0.8$ | $4.2 \pm 0.7$ | $4.6 \pm 0.8$ | <0.001 |
| Blood urea nitrogen, mmol/L | 6.0 ± 5.0 | 4.8 ± 2.3 | 12.5 ± 9.1 | <0.001 |
| Serum creatinine, μmol/L | 83 ± 84 | 68 ± 16 | 169 ± 190 | <0.001 |
| Peak serum creatinine, μmol/L | 94 ± 102 | 74 ± 38 | 200 ± 214 | <0.001 |
| eGFR, ml/min/1.73m<sup>2</sup> | 86 ± 24 | 94 ± 17 | 45 ± 16 | <0.001 |
| Proteinuria, % |  |  |  |  |
| Negative | 248/443 (56.0) | 232/389 (59.6) | 16/54 (29.6) | <0.001 |
| + | 150/443 (33.9) | 128/389 (32.9) | 22/54 (40.7) |  |
| ++~+++ | 45/443 (10.2) | 29/389 (7.5) | 16/54 (29.6) |  |
| Hematuria, % |  |  |  |  |
| Negative | 324/443 (73.1) | 299/389 (76.9) | 25/54 (46.3) | <0.001 |
| + | 68/443 (15.3) | 52/389 (13.4) | 16/54 (29.6) |  |
| ++~+++ | 51/443 (11.5) | 38/389 (9.8) | 13/54 (24.1) |  |

Data are presented as number/total (percentage) or mean ± SD; COVID-2019, coronavirus disease 2019; eGFR, estimated glomerular filtration.

### Kidney impairments

On admission, the baseline Scr was elevated in 110 (15.5%) patient. During hospitalization, the peak Scr was 94 ± 102 μmol/L. 44.0% patients had proteinuria, and relatively fewer patients (26.9%) demonstrated hematuria (Table 2).

Compared with patients with normal baseline Scr, the age and the percentage of male and severe subgroups were significantly higher in patients with elevated baseline Scr (Table 1). Moreover, patients with elevated baseline Scr demonstrated higher leukocyte count, lower lymphocyte count and platelet count. The coagulant function abnormality, including prolonged activated partial thromboplastin time and higher D-dimer, were more common in patients with elevated baseline Scr. The percentage of increased procalcitonin and the level of aspartate aminotransferase and lactose dehydrogenase were also higher in patients with elevated baseline Scr. Of note, the gap of peak and baseline Scr was much greater in patients with elevated baseline Scr (Table 2).

### Incidence of AKI and in-hospital death

During hospitalization, AKI was documented in 22 (3.2%) patients. The incidence of AKI was significantly higher in patients with elevated baseline Scr (9.1%) than patients with normal baseline Scr (2.0%). Moreover, patients with elevated baseline Scr developed more severe AKI (Table 1). Figure 1a showed serial measurement of Scr of each patient with AKI. Most of AKI occurred within 7 days after admission. For patients with normal baseline Scr, AKI occurred later and tended to recovered more rapidly. However, AKI happened more quickly and severe in patients with elevated baseline Scr.

**Figure 1.**
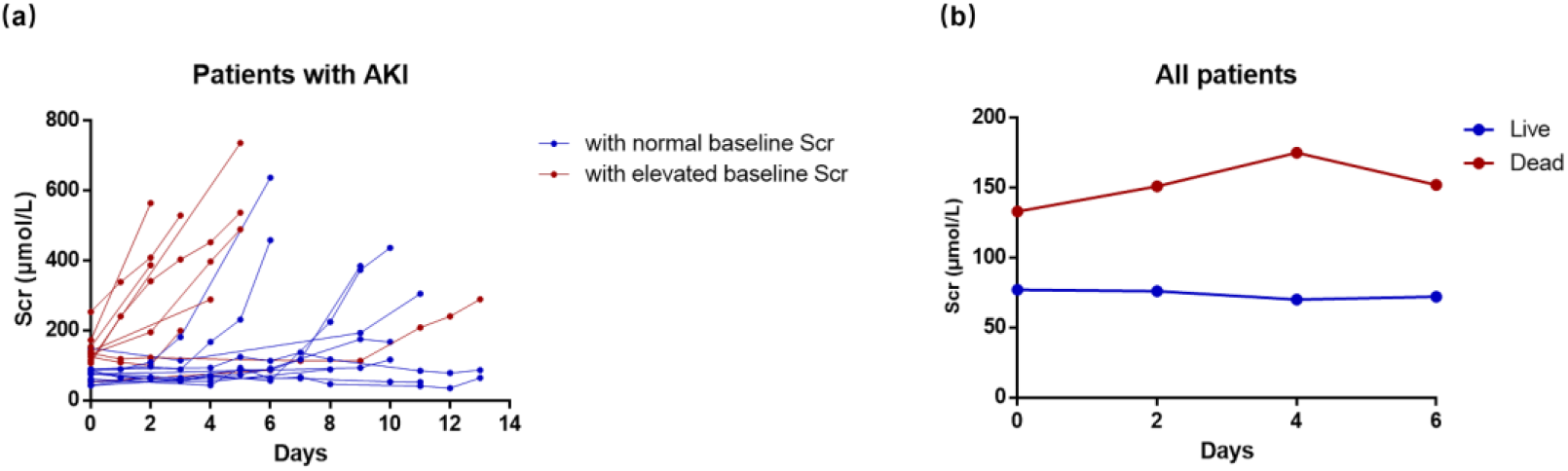
Serum creatinine changes. (a) serial measurement of serum creatinine of each patients with AKI, (b) serial measurement of serum creatinine (expressed as mean) of all patients.

The in-hospital death occurred in 89 (12.5%) patients in our study. The incidence of in-hospital death in patients with elevated baseline Scr was 30.9%, which was significantly higher than patients with normal baseline Scr (9.2%) (Table 1). The median length of stay in hospital was 5 (2-6) days for dead patients. Scr showed apparent increase in dead patients during hospitalization, while a downward trend was observed in live patients (Figure 1b).

### Association of kidney impairment indicators with in-hospital death

Kaplan-Meier analysis revealed a significantly higher in-hospital death rate for patients with kidney impairments, including elevated baseline Scr, elevated baseline blood urea nitrogen (BUN), proteinuria, hematuria and AKI (Figure 2). Univariate Cox regression analysis showed that older than age 65, severe disease, leukocyte count greater than 4×10^9^/L and lymphocyte count less than 1.5×10^9^/L were associated with in-hospital death. Besides, kidney impairment indicators mentioned above were also associated with in-hospital death (Table 3). After adjusted with age, sex, disease severity, leukocyte count and lymphocyte count, elevated baseline Scr (HR: 3.61, 95%CI: 2.18-5.98), elevated baseline BUN (HR: 2.51, 95%CI: 1.57-4.02), peak Scr > 133μmol/L (HR:2.59, 95%CI: 1.52-4.44), proteinuria (+: HR: 1.46, 95%CI: 0.56-3.81; ++∼+++: HR: 5.00, 1.89-13.21), hematuria (+: HR: 3.15, 95%CI: 1.17-8.48; ++∼+++: HR: 8.51, 3.51-20.65) and AKI (HR: 2.21, 95%CI: 1.11-4.39) were associated with in-hospital death (Table 3).

**Table 3.** Univariate Cox regression analysis of association between kidney impairment indicators and in-hospital death in patients with 2019-nCov Pneumonia Patients.

| Variables | HRs | 95%CI | P-value |
| --- | --- | --- | --- |
| Age > 65 years | 2.51 | 1.64-3.86 | <0.001 |
| Sex, male | 2.44 | 1.53-3.87 | <0.001 |
| Severe disease | 8.24 | 4.84-14.04 | <0.001 |
| Leukocyte count > 4 × 10<sup>9</sup>/L | 4.00 | 1.75-9.17 | 0.001 |
| Lymphocyte count < 1.5 × 10<sup>9</sup>/L | 5.09 | 1.25-20.68 | 0.023 |
| Elevated serum creatinine | 3.98 | 2.59-6.11 | <0.001 |
| Elevated blood urea nitrogen | 8.27 | 5.43-12.6 | <0.001 |
| Peak serum creatinine >133μmol/L | 6.04 | 3.8-9.62 | <0.001 |
| Proteinuria |  |  |  |
| Negative | reference | reference |  |
| + | 4.06 | 1.68-9.8 | 0.002 |
| ++~+++ | 11.37 | 4.54-28.51 | <0.001 |
| Hematuria |  |  |  |
| Negative | reference | reference |  |
| + | 5.47 | 2.11-14.18 | <0.001 |
| ++~+++ | 18.65 | 8.21-42.37 | <0.001 |
| Acute kidney injury | 4.92 | 2.61-9.25 | <0.001 |

**Figure 2.**
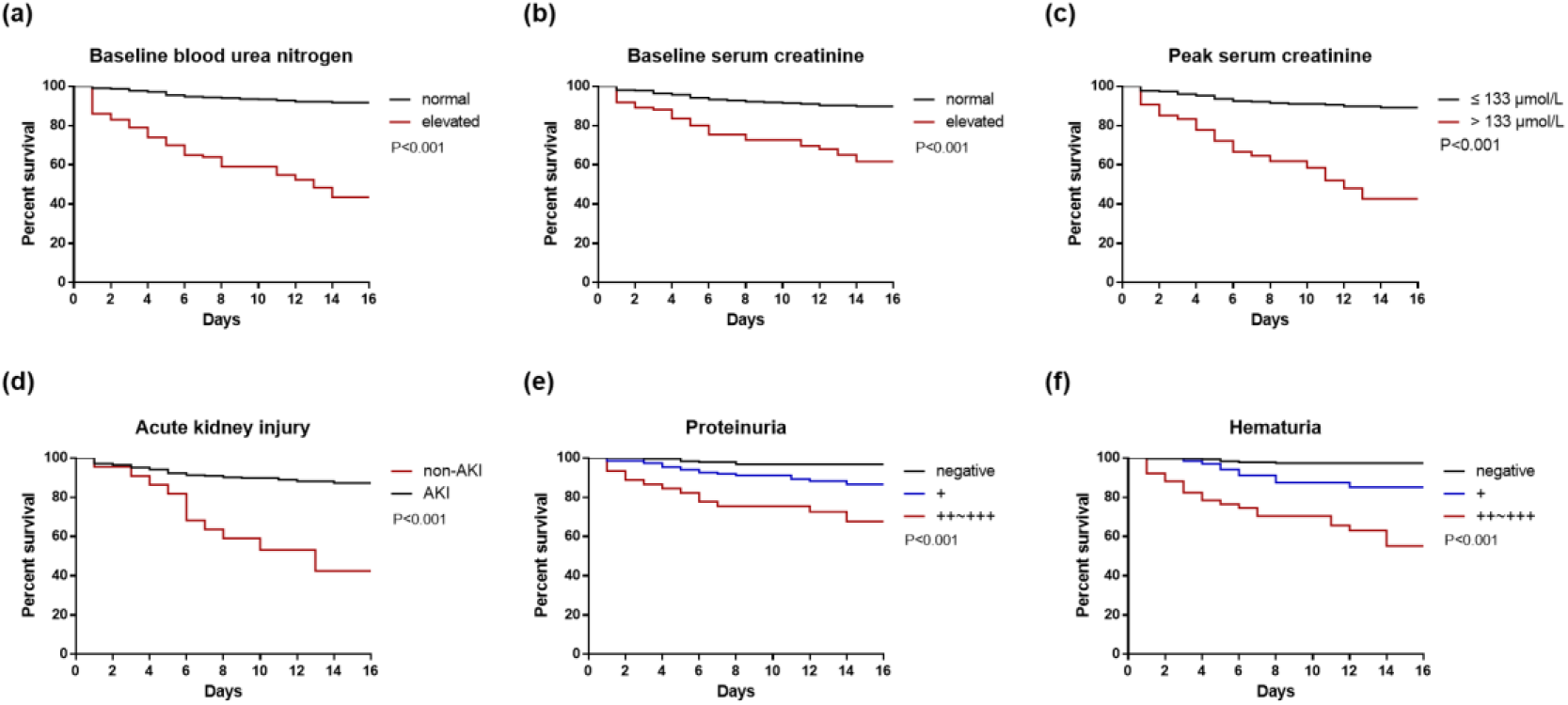
Kaplan-Meier curves for in-hospital death of patients with COVID-19 subgroup by kidney impairment indicators. (a) baseline blood urea nitrogen; (b) baseline serum creatinine; (c) peak serum creatinine; (d) acute kidney injury; (e) proteinuria; (f) hematuria.

**Figure 3.**
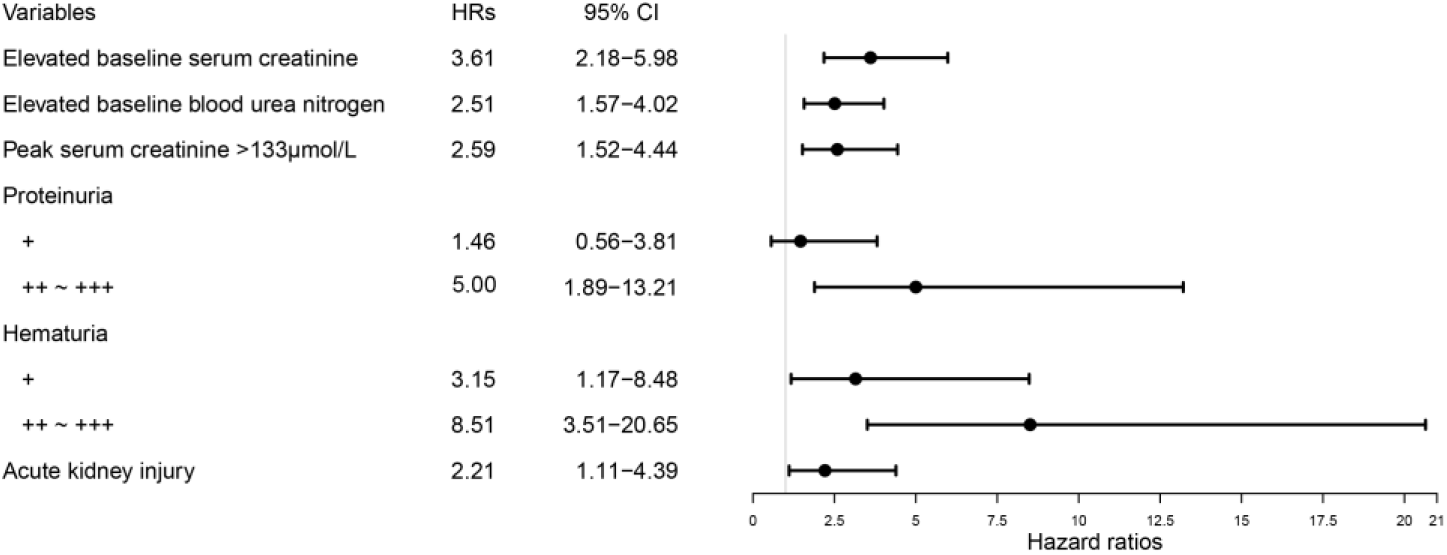
The risk of in-hospital death in patients with COVID-19. COVID-2019, coronavirus disease 2019; HRs, hazard ratios; 95%CI, 95% confidence interval; HRs of each variables was obtained with proportional hazard Cox model adjusting for age, sex, disease severity, leukocyte count and lymphocyte count.

## Discussion

In this large consecutive cohort study conducted in a tertiary teaching hospital with 3 branches in Wuhan, we have determined the prevalence of kidney impairment in hospitalized COVID-19 patients was high. By analyzing clinic data and patients’ outcome, we also demonstrated that kidney impairment was associated with in-hospital death.

In our study, a lot of patients have kidney impairment, including abnormal urinary analysis (proteinuria, hematuria) and kidney dysfunction (elevated BUN and Scr) on admission. Indeed, multiple organ involvements including the liver, gastrointestinal tract and kidney have been reported during the course of SARS in 2003^8^ and COVID-19^9^. Of note, the high prevalence of kidney impairment at baseline may be explained by the fact that many COVID-19 patients couldn’t be admitted to hospital in the early stage of disease outbreak because of large number of patients and limited beds in hospital in Wuhan. In addition, another explanation is that some COVID-19 patients may have a past history of chronic kidney disease (CKD) and CKD patients have a proinflammatory milieu and functional defects in innate and adaptive immune cell populations^10^. In a community-based cohort of nearly 10,000 adult individuals, reduced eGFR and elevated albumin creatinine ratio were associated with higher risk for hospitalization with infection and subsequent mortality^11^. Furthermore, CKD patients have a higher risk for upper respiratory tract infection^12^ and pneumonia^13^.

AKI is a syndrome of abrupt loss of kidney function that is strongly associated with higher mortality and morbidity^14^. In our cohort the detect rate of AKI in COVID-19 patients was 3.2%, which was similar to that reported in previous studies with small patients numbers^1, 4, 9, 15^ and higher than 0.5% in a large observational study^16^. This may be explained by that the proportion of severe patients was extremely high in previous case series and only 15.7% in the large observational study. In our large cohort study, 35.5% patients were severe and this may illustrate the real detection rate of AKI in clinic practice in Wuhan. Importantly, the present method of detecting AKI is mainly based on changes in Scr and the frequency of Scr tests has a substantial impact on the detection rate of AKI^17^. In a nationwide cross-sectional survey of hospitalized adult patients in China, the detection rate of AKI was only 0.99% by KDIGO criteria^18^. After adjusting for frequency of Scr, the incidence of AKI in Chinese hospitalized adults gave rise to 11.6%^19^. Thus, in order to improve early detection kidney injury, more frequent Scr measurements should be performed in the treatment of COVID-19.

The etiology of kidney impairment in COVID-19 patients is likely to be diverse and multifactorial. First, the novel coronavirus may cause direct cytopathic effect of kidney resident cells. This is supported by the detection of PCR fragments of coronavirus in blood and urine in 2003 SARS ^20^ and COVID-19 patients^4^. Recently, it was reported that the novel coronavirus uses the angiotensin converting enzyme II (ACE2) as a cell entry receptor, which is identical to that of the SARS-CoV in 2003^21^. Human tissues RNA-seq data demonstrate that the ACE2 expression in urinary organs (kidney) was much higher (nearly 100-fold) than that in respiratory organs (lung)^22^. Therefore, the kidney impairment may be caused by coronavirus entering the cells through ACE2 that are highly expressed in the kidney. Second, deposition of immune complexes of viral antigen or virus-induced specific immunological effector mechanisms (specific T lymphocyte or antibody) may damage the kidney. However, the data of kidney specimens from SARS patients showed normal glomerular histology with absence of electron-dense deposits, indicating the possibility of an active immune-mediated glomerulonephritis was low. Thus, kidney histology of COVID-19 patients is needed in the further study. Third, virus-induced cytokines or mediators have indirect effects on renal tissue, such as hypoxia, shock, rhabdomyolysis. In fact, some of the 2009 H1N1 patients showed mild to moderate elevation of creatine kinase^23^. And in 138 hospitalized patient there was an increase tendency towards an increase in creatine kinase level of COVID-19 patients in ICU^15^. Consistently, patients with kidney impairment demonstrated significant increase of creatine kinase in our study.

This is the first study indicated the association of kidney impairment and in-hospital death in patients with COVID-19. It is reported that AKI is associated with an increased risk of death in patients with SARS^5^, which is in consistence with our study. We also confirmed the value of Scr on admission in predicting the in-hospital death of patients with COVID-19 and AKI was more likely to occur in patients with elevated baseline Scr. It is noteworthy that most COVID-19 patients developed acute kidney injury in the early period of hospitalization, especially in only 2 days after admission in patients with elevated baseline Scr. Therefore, in the treatment of COVID-19, early prevention of kidney impairment, including adequate hemodynamic support and avoiding nephrotoxic drugs, is particularly important and early renal replacement treatment may improve the patients’ prognosis. Beyond insufficiency in kidney function, the abnormal urine analysis, including proteinuria and hematuria, was also associated with in-hospital death. This indicated that more attention should be paid to urine test in clinic.

Even though this study included a large number of patients from a tertiary teaching hospital in Wuhan, there are several limitations. First, this was an observational study, which may lead to underestimate of AKI or erroneous associations. However, the large number of COVID-19 case in this study may minimize the potential for bias. Second, although we attempted to adjust for many confounders, there may exist confounders either unmeasured or unknown that could explain our observed results. Third, the clinic data of patient after discharge is lacking, so we could not assess the effect of COVID-19 on long-term outcome. The further impact of COVID-19 on patients’ kidney function, especially the incidence of chronic kidney disease in these patients should be the focus of future research.

## Conclusions

The prevalence of kidney impairment (abnormal urine analysis and kidney dysfunction) in hospitalized COVID-19 patients was high. After adjustment for confounders, kidney impairment was associated with higher risk of in-hospital death. Clinicians should increase their awareness of kidney impairment in hospitalized COVID-19 patients. Early detection and effective intervention of kidney impairment may help to reduce deaths of COVID-19 patients in clinical practice.

## Data Availability

The datasets used and analysed during the current study are available from the corresponding author on reasonable request

## Abbreviations

95% CI: 95% confidence interval;
ACE2: angiotensin converting enzyme II
AKI: Acute kidney injury;
BUN: Blood urea nitrogen
CKD: Chronic kidney disease;
CKD-EPI: Chronic Kidney Disease Epidemiology Collaboration
COVID-19: Coronavirus disease 2019
eGFR: Estimated glomerular filtration rate
HR: Hazard ratio
ICU: Intensive care unit
KDIGO: Kidney Disease Improving Global Outcomes;
SARS: Severe acute respiratory syndrome
SARS-CoV-2: Severe acute respiratory syndrome coronavirus 2
Scr: Serum creatinine;
WHO: World health organization;

## Declarations

### Ethics approval and consent to participate

The study protocol and waived written informed consent was approved by the Medical Ethics Committee of Tongji Hospital (No. TJ-C20200132).

## Acknowledgements

The authors greatly appreciate all the hospital staff for their efforts in recruiting and treating patients and thank all patients involved in this study.

## Author contributions

G.X., S.G. designed the study. Y.C., R.L., K.W., M.Z., Z.W., L.D., J.L. and Y.Y. collected the data, prepared the figures and tables. Y.C. and S.G. contributed analytical tools. Y.C. and S.G. wrote the paper. S.G. and G.X. conceived the project and supervised and coordinated all the work.

